# Age-dependence of mortality from novel coronavirus disease (COVID-19) in highly exposed populations: New York transit workers and residents and Diamond Princess passengers

**DOI:** 10.1101/2020.05.14.20094847

**Authors:** Christopher T. Leffler, Matthew C. Hogan

## Abstract

**Background:** Populations heavily exposed to the novel coronavirus provide an opportunity to estimate the mortality from COVID-19 in different age groups.

**Methods:** The mortality reported by May 13 from COVID-19 among Diamond Princess cruise ship passengers, and New York residents and Metropolitan Transit Authority (MTA) workers was estimated based on publicly available information.

**Results:** The mortality among children (age 0 to 17 yrs) in New York City was 1 in 172,692. The mortality in New York state was 1 in 322,217 for ages 10-19 yrs., and 1 in 36,725 for ages 20-29 yrs. The mortality among New York transit workers was estimated to be 1 in 7,329 for ages 30-39 years; 1 in 1,075 for ages 40-49 yrs.; 1 in 343 for ages 50-59 yrs.; and 1 in 178 for ages 60-69 yrs. Among Diamond Princess passengers, the mortality was estimated to be 1 in 145 for ages 70-79, and 1 in 54 for ages 80-89.

**Conclusions:** Mortality among populations exposed to the novel coronavirus increases with age, ranging from about 1 in 170,000 below the age of 18 years, to 1 in 54 above the age of 80 years.

## Introduction

Certain confined, occupational and regional populations have had substantial exposure to the SARS-CoV-2 virus which causes the novel coronavirus disease (COVID-19). Such populations provide an opportunity to estimate the mortality which might occur after many members of a population have been exposed.

The COVID-19 outbreak occurring on the Diamond Princess cruise ship in February 2020 has resulted in the deaths of 14 passengers, and has provided a window into the effect of the virus in the elderly.^1^ In the United States, the large populations of New York state and city, the epicenter of the national outbreak, provide an opportunity to estimate mortality in children and young adults. Finally, as of May 6, 2020, 109 New York Metropolitan Transit Authority (MTA) workers have died from coronavirus.^2-4^ This population provides an opportunity to estimate the mortality in older working adults, and has not been previously studied, to our knowledge.

## Methods

We estimated from publicly available sources the coronavirus-related mortality for each age group on the Diamond Princess cruise ship, in the United States (nationally), in New York City and state, and among New York Metropolitan Transit Authority workers.^2-4^

The age breakdown of New York City’s population was determined by dividing the COVID-19 deaths by the mortality rates reported by the New York City Department of Health.^5^

The New York State population age breakdown was obtained from the census.^6^ The United States population, total and by decade, were tabulated.^7,8^

Exact binomial 95% confidence intervals for proportions were determined, unless the denominator was over 10,000, in which case Poisson confidence intervals were determined. Only data available to the public were used. The study was approved by the VCU Office of Research Subjects Protection.

## Results

### Mortality Among Younger New Yorkers

Among children in New York City (ages 0 to 17 years), the coronavirus mortality was estimated to be 1 in 173,000, with a 95% CI of 1 in 360,000 to 1 in 94,000 (Tables 1-2).^5^ Nationally, infants under age 1 tended to have a higher mortality (1 in 947,500), compared with children age 1 to 14 (1 in 9,517,000) (Table 3).^9^

**Table 1.** Mortality from novel coronavirus (COVID-19) in New York City reported by May 11, 2020.

| Age (years) | Deaths | Total in Age Group | Mortality |
| --- | --- | --- | --- |
| 0-17 | 10 | 1,726,916 | 1 in 172,692 |
| 18-44 | 596 | 3,370,166 | 1 in 5,655 |
| 45-64 | 3,383 | 2,056,112 | 1 in 608 |
| 65-74 | 3,753 | 699,112 | 1 in 186 |
| 75 and over | 7,356 | 546,976 | 1 in 74 |
| All | 15,101 | 8,398,507 | 1 in 556 |

New York City deaths reported as of May 11 at 6 PM.<sup>5</sup>

**Table 2.**
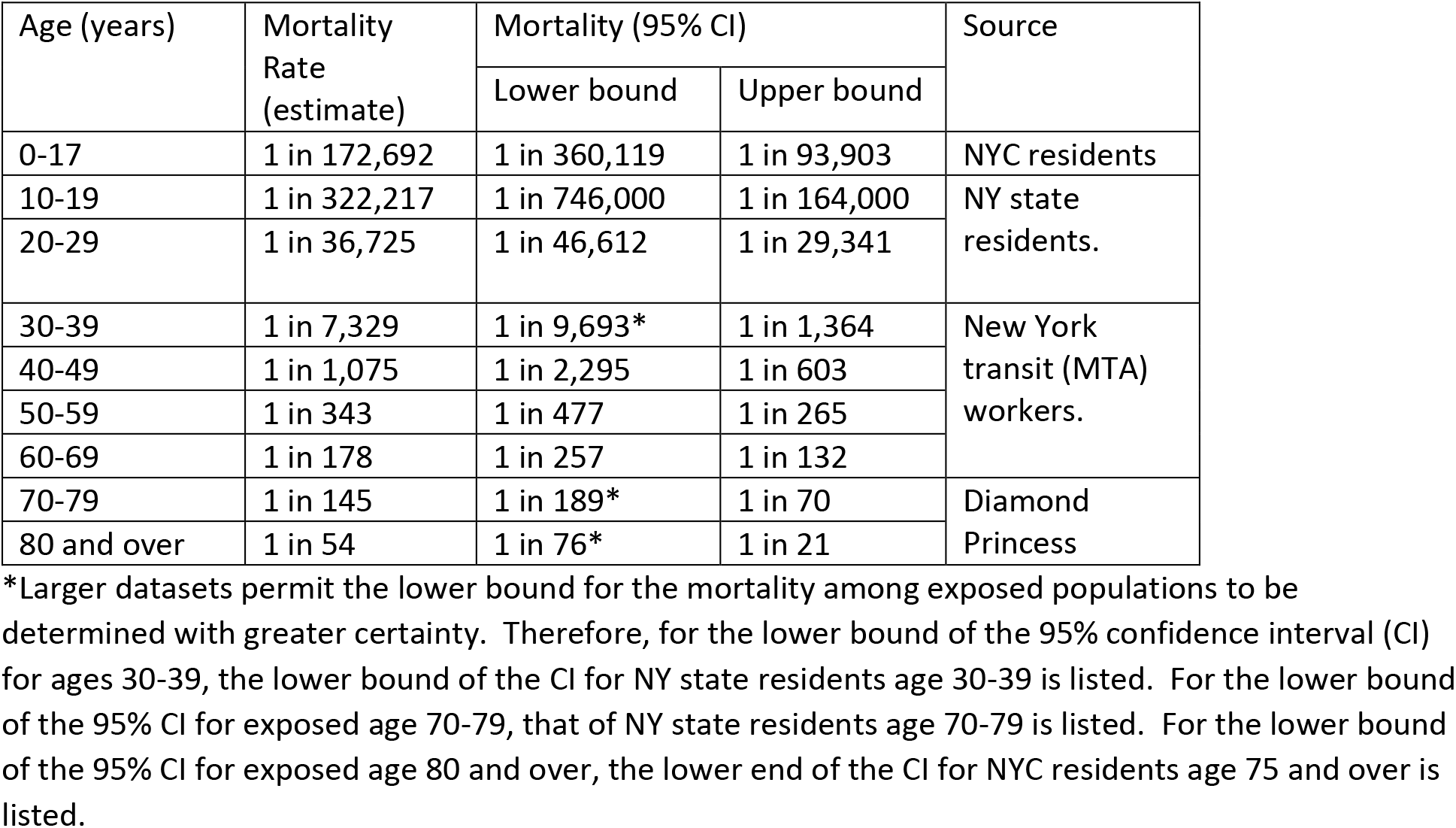
Coronavirus Mortality Among Highly-Exposed Populations Reported by May 13, 2020 (New York Residents and Transit Workers, and the Diamond Princess).

**Table 3.** Mortality from novel coronavirus (COVID-19) in the United States reported by the CDC by May 13, 2020.

| Age (years) | Deaths | Total in Age Group | Mortality |
| --- | --- | --- | --- |
| 0 (<1) | 4 | 3,790,000 | 1 in 947,500 |
| 1-4 | 2 | 16,020,000 | 1 in 8,010,000 |
| 5-14 | 4 | 41,080,000 | 1 in 10,270,000 |
| 15-24 | 48 | 42,960,000 | 1 in 895,000 |
| 25-34 | 317 | 45,690,000 | 1 in 144,000 |
| 35-44 | 796 | 41,280,000 | 1 in 51,900 |
| 45-54 | 2,262 | 41,630,000 | 1 in 18,400 |
| 55-64 | 5,422 | 42,270,000 | 1 in 7,800 |
| 65-74 | 9,359 | 30,480,000 | 1 in 3,260 |
| 75-84 | 12,026 | 15,390,000 | 1 in 1,280 |
| 85 and over | 13,776 | 6,550,000 | 1 in 475 |
| All | 44,016 | 330,709,369 | 1 in 7,510 |

United States coronavirus deaths reported to the CDC, from Feb 1, 2020 to May 6, 2020. This was the latest available information on the CDC website on May 13, 2020.<sup>9</sup>

In New York State, the coronavirus mortality reported by May 13 was estimated as 1 in 322,217 for ages 10-19 years; 1 in 36,725 for ages 20-29 years; and 1 in 8,617 for ages 30-39 years (Tables 2, 4).^10^

**Table 4.** Mortality from Novel Coronavirus in New York State by May 13, 2020.

| Age (years) | Deaths | Total in Age Group | Mortality |
| --- | --- | --- | --- |
| 0-9 | 4 | 2,319,777 | 1 in 579,944 |
| 10-19 | 8 | 2,577,734 | 1 in 322,217 |
| 20-29 | 76 | 2,791,112 | 1 in 36,725 |
| 30-39 | 294 | 2,533,284 | 1 in 8,617 |
| 40-49 | 788 | 2,814,656 | 1 in 3,572 |
| 50-59 | 2,122 | 2,657,336 | 1 in 1,252 |
| 60-69 | 4,321 | 1,839,471 | 1 in 425 |
| 70-79 | 5,768 | 1,062,198 | 1 in 184 |
| 80 and over | 8623 | 782,534 | 1 in 91 |
| Total | 22,013 | 19,378,102 | 1 in 880 |

New York State coronavirus deaths reported by May 13, 2020.<sup>10</sup>

### Adult Mortality and New York City Transit

The New York MTA has 74,000 employees, most of whom are represented by the Transport Workers Union.^11^ In total, 106 of the 109 MTA workers who had died of coronavirus as of May 6, 2020 were in the subways and buses division, which has 55,000 employees.^12^

Of the 68 New York City MTA workers who died of coronavirus, and whose ages could be determined from public sources, one was in his 30s (1.5%, 95% CI 0.04-7.9%), 9 were in their 40s (13.2%, 95% CI 6.2-23.6%), 29 were in their 50s (42.7%, 95% CI 30.7-55.2%), 25 were in their 60s (36.8%, 95% CI 25.4-49.3%), and 4 were in their 70s (5.9%, 95% CI 1.6-14.4%). None was younger than 39 or older than 73. The age-distribution of known deaths was applied to the 106 total deaths in the bus and subways division to estimate the numbers of deaths in each decade (Table 5).^2-4,13^

**Table 5.** Mortality from Novel Coronavirus among New York MTA (Transit) Workers by May 6, 2020.^2-4,13^

| Age (years) | Estimated Deaths | Total in Age Group | Mortality |
| --- | --- | --- | --- |
| 18-19 | 0 | 240 | -- |
| 20-29 | 0 | 4,240 | -- |
| 30-39 | 1-2 | 11,425 | 1 in 7,329 |
| 40-49 | 14 | 15,080 | 1 in 1,075 |
| 50-59 | 45 | 15,520 | 1 in 343 |
| 60-69 | 39 | 6,931 | 1 in 178 |
| 70-79 | 6 | 1,274 | 1 in 204 |
| Total | 106 | 55,000 | 1 in 519 |

The age distribution of MTA employees is: 18-25 years (1.7%), 26-35 years (16.0%), 36-54 years (53.1%), 55-62 years (22.8%), and over 63 years (6.4%).^13^ From this distribution, the number of the 55,000 bus and subway workers was calculated for each age decade (Table 5).

Among New York MTA workers during the pandemic, the mortality was estimated to be 1 in 7,329 for ages 30-39; 1 in 1,075 for ages 40-49; 1 in 343 for ages 50-59; 1 in 178 for ages 60-69 (Tables 2, 5).

### The Diamond Princess and Elderly Mortality

The age ranges of the passengers on the Diamond Princess cruise ship at the time of the coronavirus outbreak have been published (Table 6).^1^ A total of 14 of the 3711 passengers have died from coronavirus, as of May 13, 2020, including one woman in her 60s (Table 6).^14^

**Table 6.**
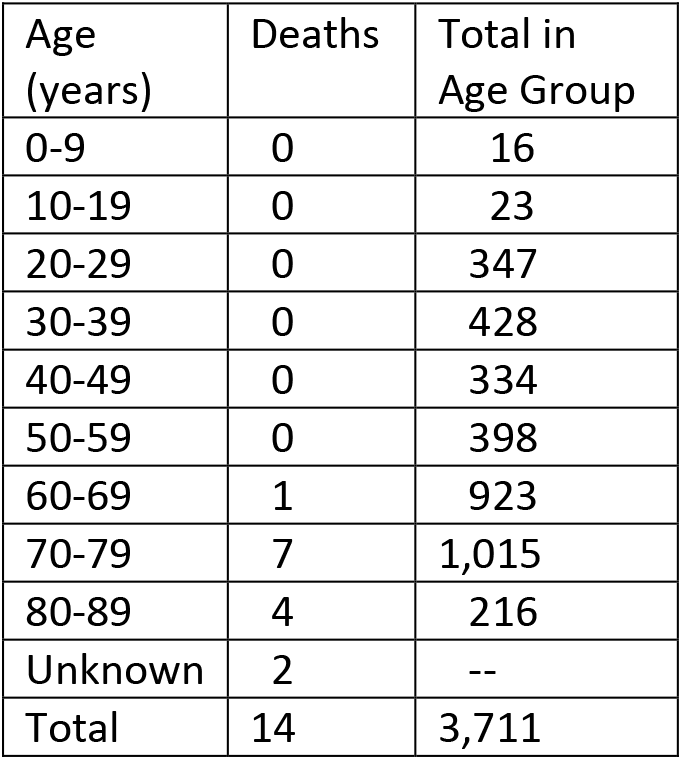
Mortality from Novel Coronavirus Among Diamond Princess Passengers.^1,14-24^

| Age (years) | Deaths | Total in Age Group |
| --- | --- | --- |
| 0-9 | 0 | 16 |
| 10-19 | 0 | 23 |
| 20-29 | 0 | 347 |
| 30-39 | 0 | 428 |
| 40-49 | 0 | 334 |
| 50-59 | 0 | 398 |
| 60-69 | 1 | 923 |
| 70-79 | 7 | 1,015 |
| 80-89 | 4 | 216 |
| Unknown | 2 | -- |
| Total | 14 | 3,711 |

Seven Diamond Princess passengers in their 70s died of COVID-19.^15-20^ For ages 70 to 79 years, the mortality was 1 in 145, with 95% confidence limits from 1 in 357, to 1 in 70 (Tables 2, 6). A lower bound for mortality in exposed populations from 70 to 79 years of age comes from the data of New York State, where the mortality was 1 in 184, with 95% confidence limits of 1 in 189 to 1 in 179 (Tables 2, 4).

Four Diamond Princess passengers in their 80s died of COVID-19.^21-22^ The mortality was for this age group was 1 in 54, (95% CI 1 in 196, to 1 in 21, Tables 2, 6). A lower bound for mortality in the exposed elderly is obtained from the data of New York City, where the observed mortality for ages 75 and over was 1 in 74, with 95% confidence limits of 1 in 76, to 1 in 72 (Tables 1-2).

The mortality for these age decades on the Diamond Princess might actually be higher, but the ages of 2 of the 14 passengers who died was not disclosed (Table 6).^23,24^

## Discussion

This study found that the mortality from coronavirus among highly exposed populations is heavily dependent on age, ranging from about 1 in 170,000 under age 18, to 1 in 54 for those in their 80s. It will be important for all people, especially older adults, to take precautions to avoid infection. The strength of this study is that our method did not require estimation of the prevalence of infection or case fatality ratios (since there is no consensus about these values yet). It should be emphasized that the tabulated mortality rates are *not* infection fatality rates (fractions of infected people who die). Rather, the listed mortality rates are the fraction of the entire population who died.

Children seem to be at a low risk of mortality from coronavirus, around 1 in 170,000 in New York City to date. However, the national data do suggest that infants may be at a higher risk of coronavirus-related mortality than other children. As schools have been closed during the pandemic, the exposure of children to the virus, even in New York, was in all likelihood less than that of the other groups studied (MTA workers, Diamond Princess passengers). Presumably, if schools reopen, the mortality among children will increase. Still, it is likely that the mortality among children will remain several orders of magnitude below that of older adults.

The mortality among young adults age 20-39 is fairly low, but still an important consideration for both individual and institutional decision-making. One New York MTA worker in his late 30s died of coronavirus. A second New York transit worker in his mid-30s died of coronavirus, though technically he was not an MTA employee, and therefore, did not factor in our analysis.^2^ The New York state data confirms that these deaths of workers in their 30s were not unprecedented. It would be useful if localities (and national authorities) could release coronavirus demographic data broken down with more granularity. For instance, colleges would be most interested in the data from age 18-22 years.

This was the first study of the mortality among New York transit workers, to our knowledge. Analysis of this group helps to characterize the mortality among older working age adults. The first confirmed case of coronavirus infection in New York City was reported on March 1, 2020.^25^ The MTA workers were forbidden to wear masks and gloves until March 8.^26,27^ On March 26, two MTA workers died from coronavirus—the first MTA workers to suffer this fate.^28^ An announcement that the MTA would provide the workers with masks on came on March 27.^29^ On April 4, 2020, the MTA was able to obtain 250,000 N95 masks for the workers.^30^ On April 15, the governor of New York announced an order to require that after a 3-day notification period, the public would be required to wear masks when unable to socially distance, such as on public transit.^31^ Therefore, there was a delay of 5 weeks between presence of the infection in the city and the MTA obtaining masks, and of 7 weeks until the public was required to wear masks when in transit.

Based on the New York transit worker data, the mortality risk among exposed populations continues to increase progressively in the 40s and 50s. The sole fatality on the USS Theodore Roosevelt, on which a coronavirus outbreak occurred, was a 41-year-old sailor.^32^

In highly-exposed populations, the fraction of the population infected has been less than 100%. On some ships with outbreaks, all on board were tested. On the USS Theodore Roosevelt, 856 of 4845 sailors (17.7%) tested positive for the virus.^32^ On the Diamond Princess, 619 of 3,711 passengers (16.7%) tested positive for coronavirus, only half of whom were symptomatic.^1^ Among New York MTA workers, a group which was not universally tested, over 6000 workers (8%) have either tested positive (over 2000), or entered quarantine (4000 workers).^33^ In these populations, it is not known if those who tested negative were false negatives, were tested at the wrong time, had less exposure than their fellow passengers or workers, or were less susceptible to infection.

This study has a number of limitations. If the Diamond Princess passengers or MTA workers had not all been exposed to the virus, then the observed mortality underestimates the potential mortality. Also, there might be differences between the Diamond Princess population or the MTA workers and the population at large. The MTA workers were mostly minorities, and one could argue that they suffered excess mortality because of preexisting medical conditions. On the other hand, many of the most disabled are excluded from working or travelling at sea. Our study did not estimate the mortality among nursing home patients, which has been substantially higher than that of the general population.

## Data Availability

Data are available from the author.

